# Scenario modelling for diminished influenza seasons during 2020/2021 and 2021/2022 in England

**DOI:** 10.1101/2022.10.27.22281628

**Authors:** Edward M. Hill, Matt J. Keeling

## Abstract

**Background:** There has been concern throughout the COVID-19 pandemic over highly stressed healthcare capacities being further pressured by seasonal influenza epidemics. Interventions to tackle the spread of SARS-CoV-2, the causative agent of COVID-19, have unsettled the respiratory pathogen landscape, including worldwide patterns of influenza activity. The implications of these disruptions for subsequent influenza seasons has been uncertain.

**Methods:** To conduct scenario analyses ahead of the 2020/2021 and 2021/2022 influenza seasons in England, we used a pre-existing age-structured, multi-strain compartmental model of influenza transmission and case severity, which included propagation of immunity between influenza seasons and had been previously fit to historical data. For the pre-2020/2021 influenza season, our scenarios varied the level of vaccine uptake and the inclusion/exclusion of nonpharmaceutical interventions (NPIs). We estimated the relative amount of health episode occurrences: symptomatic cases resulting in a GP consultation, hospital inpatient admissions, fatalities. In the pre-2021/2022 influenza season analysis, compared with a counterfactual case where influenza activity remained at historic levels in the 2020/2021 influenza season, we estimated the change in the same set of health episode occurrences in the 2021/2022 influenza season when assuming there was no influenza in circulation during the 2020/2021 influenza season.

**Results:** Attaining coverage of 75% in target groups for the 2020/2021 influenza season reduced health episode occurrences by 40–50% when compared to maintaining the 2019/2020 vaccination programme coverage and uptake levels. Having NPIs maintained throughout the entire influenza season saw 60-80% reductions in severe case outcomes. Combining an expanded vaccination programme and the use of NPIs could suppress the seasonal influenza epidemic, with reductions as much as 90–100%. In the absence of influenza transmission during the 2020/2021 influenza season, under our modelling assumption of mixing patterns returning to pre-2020 levels we projected a compensatory influenza epidemic with 1.2 to 2.2 times as many severe health episode occurrences in the subsequent 2021/2022 influenza season.

**Conclusions:** In the context of the time the work was originally conducted, the modelled scenarios indicated how bolstering vaccine coverage and reduction in contacts could likely allay resurgent seasonal influenza epidemics. Our analyses of the winter pressures that may be inflicted by other respiratory infections during the COVID-19 pandemic are one example of the modelling insights provided to the Scientific Pandemic Influenza Group on Modelling, Operational sub-group (SPI-M-O) for the Scientific Advisory Group for Emergencies (SAGE) in the UK.

## 1 Introduction

The public health burden imposed by seasonal influenza epidemics is variable. Different influenza types and strains produce different effects in terms of the balance between primary and secondary care pressure, which is partly related to severity by age, as well as vaccine match and uptake. There have been concerns of the potential additional burden faced in the event of a “twindemic”, where the typical seasonal influenza epidemics occur on top of COVID-19 pressures, applying further stresses on already overwhelmed healthcare services [1]. Hence, heading into the winters of 2020 to 2021 and 2021 to 2022, respectively, there were worries that if the influenza season were to have a substantial epidemic, it would produce a challenging scenario of limited healthcare capacities being put under further stress, with consequential additional increases in morbidity and mortality.

During both the 2020/2021 and 2021/2022 winter seasons COVID-19 case rates were high. In an attempt to counter the transmission of SARS-CoV-2 and curb resultant COVID-19 case burdens, NPIs such as social distancing and wearing face coverings were used. Literature began to arise throughout 2020 on the knock-on impacts of these SARS-CoV-2 targeted public health measures on the transmission of seasonal influenza amongst populations. There was evidence from Hong Kong of SARS-CoV-2 targeted NPIs also suppressing transmission of influenza [2]. National influenza laboratory surveillance data from Scandinavia demonstrated a decline in transmission of influenza and the ending of the 2019/2020 influenza season earlier than expected [3]. Furthermore, empirical observations from temperate zones in the Southern Hemisphere had indicated the absence of their typical influenza season. Very few influenza detections had been reported, despite continued or even increased testing for influenza in some countries [4, 5].

That being said, prior to September 2020 there remained uncertainty on the impact of NPIs on influenza transmission, in part due to the role of children in transmission and the altered immunity landscape due to previous influenza vaccinations administered via the national seasonal influenza vaccination programme. Other actions were also being taken to bolster the armoury of interventions against seasonal influenza, namely an expansion of seasonal influenza vaccination programmes [6]. In England, the seasonal influenza vaccination is available every year on the NHS for free to specific target groups, to help protect adults and children at risk from influenza and its complications. Ahead of the 2020/2021 influenza season, the influenza vaccine was freely offered to those aged 50–64 years old for the first time and the schools programme was expanded to include all secondary school pupils (up to and including Year 11) [7].

In this study, we detail our use of a previously formulated mathematical model describing seasonal influenza transmission and case burden in England to perform scenario analyses that scrutinised bespoke questions ahead of the 2020/2021 and 2021/2022 influenza seasons, respectively. Prior to the 2020/2021 influenza season, our scenarios studied the implications of modification to contact patterns (due to use of NPIs) and vaccination coverage on the anticipated seasonal influenza epidemics for the 2020/2021 winter season. Ultimately, with the UK experiencing a very mild 2020/2021 influenza season that had low numbers of confirmed influenza cases reported by surveillance systems [8], heading into the 2021/2022 influenza season the prominent question that we then studied was how a year of influenza transmission absence may impact seasonal influenza epidemiological outcomes.

The work described in this manuscript was presented to the Scientific Pandemic Influenza Group on Modelling, Operational sub-group (SPI-M-O) for the Scientific Advisory Group for Emergencies (SAGE) in the UK. The work for the 2021/2022 influenza season has also featured as part of a Welsh Government Technical Advisory Group report [9].

## 2 Methods

We present model results for a collection of scenarios for the 2020/2021 influenza season (work originally conducted in September 2020) and the 2021/2022 influenza season (work originally conducted in June 2021), performed using a pre-existing seasonal influenza transmission and case severity model for England [10].

In the Methods we provide: (i) a summary of the seasonal influenza epidemiological model (Section 2.1), (ii) an overview of the assumptions associated with the implementation of vaccination (Section 2.2), (iii) the simulation protocol used to assess the 2020/2021 influenza season scenarios (Section 2.3), (iv) the simulation protocol used to assess the 2021/2022 influenza season scenarios (Section 2.4), and (v) a description of the epidemiological outcomes outputted and analysed (Section 2.5).

### 2.1 Seasonal influenza epidemiological model

Herein we give a high-level summary of the constituent components of the seasonal influenza epidemiological model.

For a complete description, we refer the reader to Hill *et al*. [10]. We also provide additional details on the model framework in the Supporting Information.

#### Disease state structure

The model formulation was a system of age-structured, multi-strain SEIR-type compartmental ODEs, which had been calibrated to primary care consultations for influenza attributed GP consultations in England for the influenza seasons 2012/2013 to 2017/2018 inclusive [10].

At this juncture, we note that the model framework does not include healthcare workers or care-homes, which can be particularly affected by influenza.

#### Susceptibility

We used an age-dependent (and strain independent) susceptibility profile with age ranges (in years) of: 0–17, 18–64, 65–84, 85+. We assumed susceptibility to be homogeneous within each age band.

#### Immunity propagation

The age- and strain-structured mathematical model for seasonal influenza transmission dynamics also incorporated mechanisms for immunity propagation.

In brief, it included three susceptibility modifying factors: (i) modified susceptibility to strain *m* given infection by a strain *m* type virus the previous influenza season; (ii) carry over cross-reactivity protection between influenza B lineages; (iii) residual strain-specific protection carried over from the prior season influenza vaccine we let the immunity propagation due to vaccination in the previous season be linearly tied to the age- and strain-specific vaccine efficacy in the previous season.

We parameterised our immunity propagation mechanisms by sampling from the posterior distributions inferred when calibrating the model to influenza attributed GP consultations in England for the influenza seasons 2012/2013 to 2017/2018 inclusive. We expect alternative parameterisations of the immunity propagation processes to result in notably different outcomes; with no propagation of immunity (resulting from either natural infection or vaccination), we would not anticipate any difference between the scenarios, whereas we envision a heightened immunity propagation would increase the relative differences between the scenarios.

### 2.2 Seasonal influenza vaccination assumptions

We next expand on the assumptions applied for the uptake and effectiveness of the seasonal influenza vaccination.

#### Action of vaccination

We assumed a ‘leaky’ vaccine, where groups that received the vaccine had reduced susceptibility in line with the vaccine efficacy against each particular strain. Vaccinated individuals that became infected had an unmodified transmission potential. In other words, the infectiousness of an infected, vaccinated individual was the same as the infectiousness of an infected, unvaccinated individual.

#### Vaccine uptake

For influenza seasons prior to 2020/2021, we used the recorded uptake data from the respective influenza season.

For the vaccine uptake assumptions applied in our scenarios conducted prior to the 2020/2021 influenza season, see Section 2.3. For the vaccine uptake assumptions applied in our scenarios conducted prior to the 2021/2022 influenza season, see Section 2.4.

#### Vaccine efficacy

For influenza seasons prior to 2020/2021, we used central vaccine efficacy estimates stratified by age and influenza type, strain or lineage (where available). Other assumptions based on historical efficacy values could be implemented and would generate considerably more variation in the results.

For the 2020/2021 and 2021/2022 influenza seasons, we assumed a 50% vaccine efficacy towards each strain.

### 2.3 Simulation outline: Pre-2020/2021 influenza season analysis

#### Scenarios

We considered four scenarios in our assessment conducted prior to the 2020/2021 influenza season, reflecting different vaccination coverage and contact pattern assumption combinations.

#### Baseline (non-COVID scenario)

For the 2020/2021 influenza season vaccine coverage, we assumed 2019/2020 influenza season vaccine target groups (at-risk individuals of any age, all those aged 2, 3 or 65 years and above, and schools years reception-year 7 inclusive), uptake coverage and uptake temporal profiles. Contact patterns were unamended.

#### Expanded vaccination programme only

Vaccination for the 2020/2021 influenza season was as described in the subsection *Expanded vaccination programme for the 2020/2021 influenza season* below. Contact patterns were unamended.

#### Usage of NPIs

Vaccination for the 2020/2021 influenza season was as described in the baseline scenario. Contact patterns were modified as described in the subsection *Contact patterns in the presence of NPIs*.

#### Combination of NPIs and expanded vaccination programme

Vaccination for the 2020/2021 influenza season was as described in the subsection *Expanded vaccination programme for the 2020/2021 influenza season* below. Contact patterns were modified as described in the subsection *Contact patterns in the presence of NPIs*.

We ran 100 simulations per scenario, from the start of the 2009/2010 influenza season (i.e. September 2009) up to and including the 2020/2021 influenza season. In each realisation we sampled parameters from their respective posterior distributions from the fitted model. We performed age-stratified comparisons to the baseline scenario for the following age-brackets (in years): 0–1; 2–17; 18–49; 50–64; 65–84; 85+.

#### Expanded vaccination programme for the 2020/2021 influenza season

The plan for the national influenza immunisation programme in England for 2020 to 2021 included offering the vaccine for free on the NHS to at-risk individuals of any age, all those aged 2, 3 or 50 years and above, and schools years reception-year 7 inclusive [7].

We modelled 75% coverage for the previously stated target groups. For at-risk individuals of any age, all those aged 2, 3 or 65 years and above, and schools years reception-year 7 inclusive, we set the temporal profile for uptake in these groups to match historically observed rates.

As the 50-64 year old age group were being added to the vaccination programme for the first time in 2020 to 2021, we had no historical data to inform a temporal uptake profile. Consequently, we assumed a constant daily uptake rate throughout November 2020 - January 2021.

#### Contact patterns in the presence of NPIs

We used estimates of contacts relative to a pre-pandemic setting for England, sampled from posterior distributions of regional contact pattern estimates inferred by the Warwick SARS-CoV-2 transmission ODE model [11]. To produce a national estimate, we multiplied the regional estimates from the Warwick SARS-CoV-2 transmission ODE model by the ratio of the population in that region for that age band, divided by the total UK population for that age band. We then took the median over the posteriors to give the amended contact structure used in our active NPIs scenario.

### 2.4 Simulation outline: Pre-2021/2022 influenza season analysis

#### Scenarios

We considered two scenarios in our pre-2021/202 influenza season analysis. These two scenarios both began from the start of the 2009/2010 influenza season (i.e. September 2009), and were run up to and including the 2021/2022 influenza season:

1. **Counterfactual (no SARS-CoV-2 scenario)** No perturbations applied to influenza transmission during the time horizon of the simulations.
2. **SARS-CoV-2 impacted scenario:** Modified transmission by assuming no influenza transmission occurred during the 2020/2021 influenza season.

As for the scenarios run for the 2020/2021 influenza season, we carried out 100 simulations per scenario, sampling from the posterior distributions of the fitted transmission model in each realisation.

Once again, we performed age-stratified comparisons to the baseline scenario for the following age-brackets (in years): 0–1; 2–17; 18–49; 50–64; 65–84; 85+.

#### Vaccine uptake assumptions for the 2021/2022 influenza season

For both the counterfactual and SARS-CoV-2 impacted scenarios, for the 2021/2022 influenza season scenarios we assumed similar overall uptake and temporal profiles for uptake compared to the 2020/2021 influenza season (now these data were available as of June 2021, when this particular piece of modelling work was conducted) for the target groups: at-risk individuals of any age, all those aged 2, 3 or 65 years and above, and schools years reception-year 7 inclusive.

For the 50-64 year old age group, we assumed a constant daily uptake rate throughout November 2021 - January 2022.

Note that subsequent to us performing this analysis in late June 2021, it was announced that the influenza vaccination programme would be extended in the 2021/2022 influenza season to four additional cohorts in secondary school; all those from years 7 (11-12 year olds) to year 11 (15-16 year olds) would be offered vaccination [12].

With our interest being in relative comparisons between the modelled outputs for the two scenarios, we would not expect the qualitative conclusions to alter were we to have included vaccination of all secondary school year groups for the 2021/2022 influenza season.

### 2.5 Epidemiological outcomes

For the pair of analyses, we present comparisons relative to the counterfactual scenario for four health episode outcomes: (i) symptomatic cases that would have resulted in a GP consultation (pre-COVID restrictions); (ii) inpatient admissions; (iii) in-hospital deaths; (iv) out of hospital deaths.

In addition to evaluating these outcomes for the overall population, we produced outputs for the low-risk and at-risk groups.

We determined severe clinical outcomes by scaling cases by age- and type-specific hospitalisation and mortality ratios. We expand on our case severity assumptions in the Supporting Information.

We performed all model simulations in Julia v1.5. The system of ODEs was solved numerically with the package DifferentialEquations v6.8.0. We generated the figures in Matlab R2022a. The code repository for the study is available on GitHub: https://github.com/WarwickSBIDER/diminished-influenza-seasons.

## 3 Results

### 3.1 Scenarios conduced prior to the 2020/2021 influenza season

For the expanded vaccination scenario, attaining coverage of 75% in target groups and in 50-64 year olds for the 2020/2021 influenza season reduced health episode occurrences by 40-50% when compared to maintaining the 2019/2020 vaccination programme uptake levels (Fig. 1, left hand group of violins in each panel). We saw greater reductions in health episode occurrences for 50-64 year olds due to their novel inclusion in the vaccination programme. For those at-risk, we observed marginally greater reductions in outcome measures for those aged 0–1, 18–49 and 50–64 (Fig. S1(a)). For those of low-risk status, the expanded vaccination scenario, including 75% uptake across all 50–64 years olds, resulted in the observed starker decrease in the relative amount of clinical events of those ages compared to other age groups (Fig. S1(b)).

**Fig. 1:**
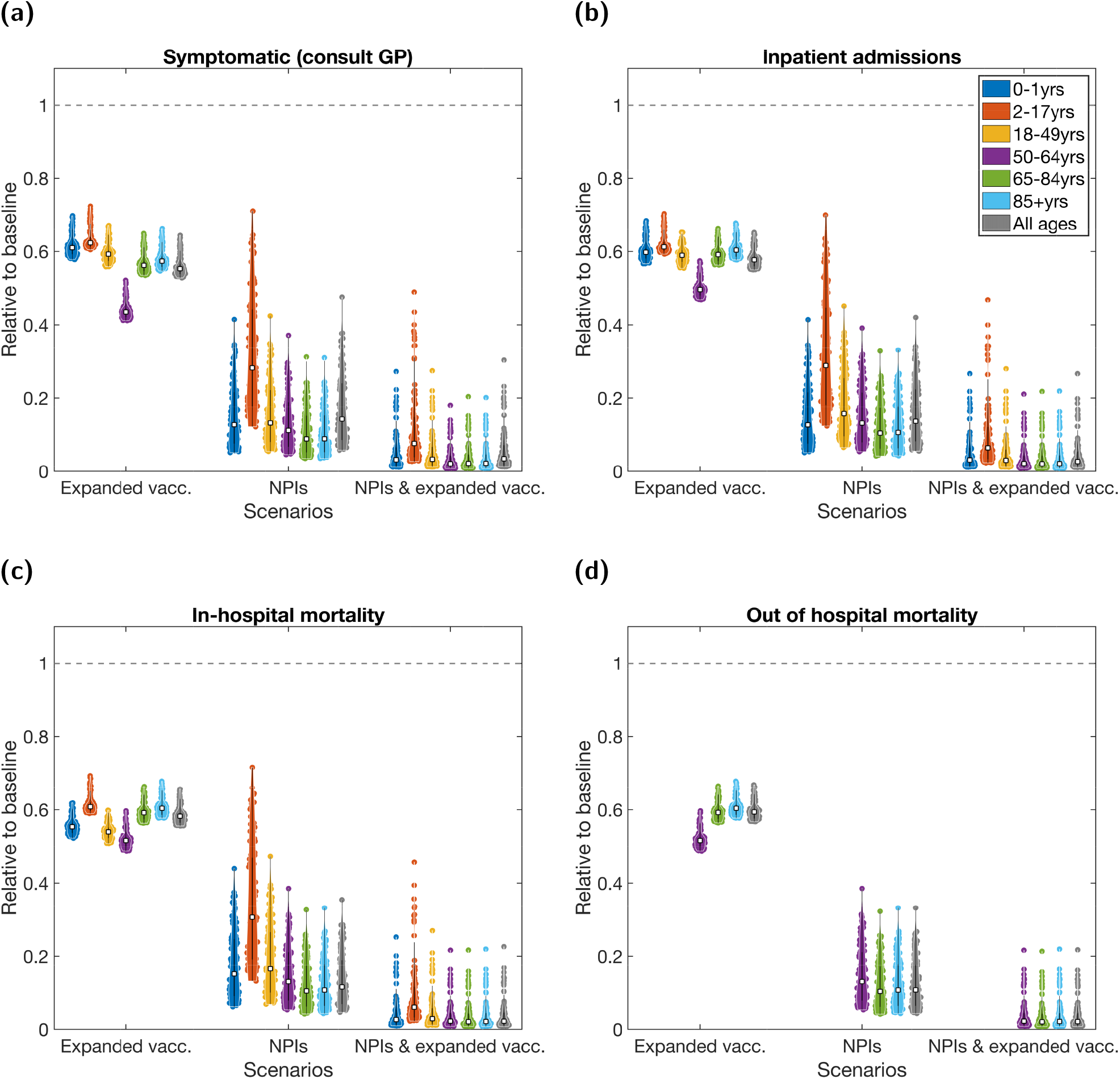
For the 2020/2021 influenza season scenarios, the age-stratified health episode occurrences relative to the baseline scenario. For each scenario, each age grouping is measured relative to the outcomes attained for that age grouping under the baseline scenario. We present relative occurrences for: **(a)** symptomatic cases resulting in a GP consultation; **(b)** hospital inpatient admissions; **(c)** in hospital fatalities; **(d)** out of hospital fatalities. In each panel, we batch the violin plots into three groups, corresponding to one of the three non-baseline scenarios: the left group for the expanded vaccination scenario, the central group for the NPIs scenario and the right group for the combined NPIs and expanded vaccination scenario. In all panels, white squares represent the medians and solid black lines the interquartile range. These results were produced accounting for both low- and at-risk groups. See Table S3 for median values and 95% prediction intervals.

Given a scenario of NPIs being maintained throughout the entire influenza season, for the majority of age groups we found an 80% reduction in severe case outcomes, with 2–17 years olds having a lesser reduction in the region of 60–80% (Fig. 1, central group of violins in each panel); the extent of this difference was likely affected by our assumptions regarding school reopening effects.

The combined usage of NPIs and an expanded vaccination programme could suppress the seasonal influenza epidemic, with reductions as much as 90–100% (Fig. 1, right hand group of violins in each panel). Under these scenarios, there were similar quantitative outcomes for both the at-risk and low-risk portions of the population (Fig. S3).

### 3.2 Scenarios conduced prior to the 2021/2022 influenza season

Here, we present comparisons relative to the counterfactual (no SARS-CoV-2) scenario for the four health episode outcomes: symptomatic (consult GP), inpatient admissions, in-hospital deaths, out of hospital deaths.

Relative to the counterfactual scenario (seasonal influenza transmission having not been perturbed during the previous influenza season), models simulations indicated it being almost certain that having no influenza transmission during the previous influenza season, 2020/2021 in our context would result in a larger burden of cases, hospitalisations and deaths in the next influenza season, 2021/2022 in our context (Fig. 2). Stratifying by the at-risk and low-risk populations, we obtained similar results for the amount of health episode occurrences relative to the counterfactual scenario (Fig. S4).

**Fig. 2:**
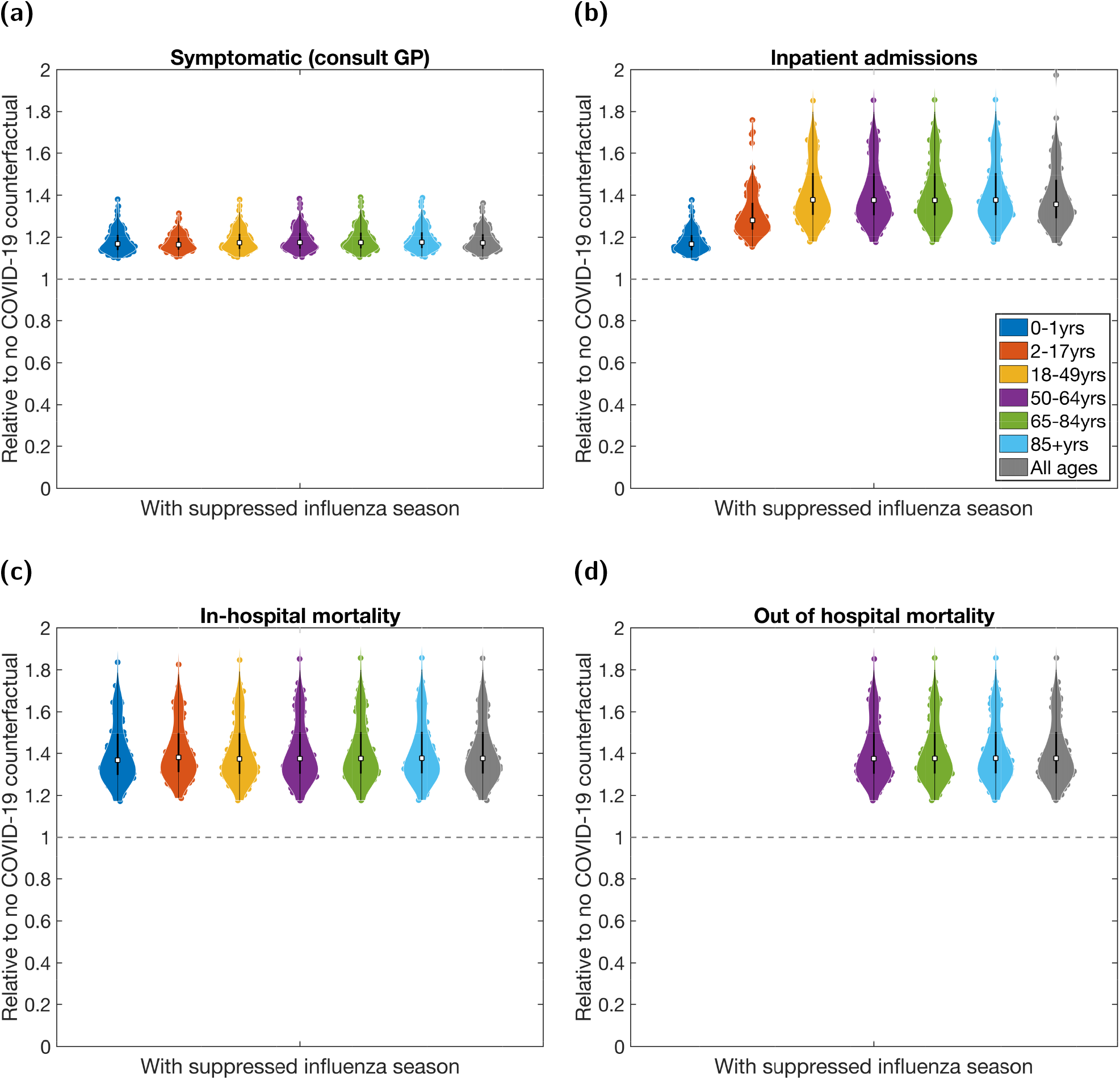
For the 2021/2022 influenza season scenarios, the age-stratified health episode occurrences relative to the non-COVID counterfactual scenario. Each age grouping is measured relative to the outcomes attained for that age grouping under the counterfactual scenario. We present relative occurrences for: **(a)** symptomatic cases resulting in a GP consultation; **(b)** hospital inpatient admissions; **(c)** in hospital fatalities; **(d)** out of hospital fatalities. In all panels, white squares represent the medians and solid black lines the interquartile range. These results were produced accounting for both low- and at-risk groups. See Table S4 for median values and 95% prediction intervals.

Across the considered epidemiological outcomes we observed broader ranges in the relative amounts of hospitalisations and deaths than in symptomatic cases resulting in GP consultation. However, these outcomes arose due to the multi-strain dynamics and the modelling assumptions for the mapping of cases to severe outcomes for the two influenza types (type A and type B); in particular, the proportion of cases attributable to type B in the model simulated 2021/2022 influenza season was typically higher in the counterfactual scenario than the SARS-CoV-2 impacted scenario (Fig. 3). We expect having alterations to the strain composition would result in notably different outcomes.

**Fig. 3:**
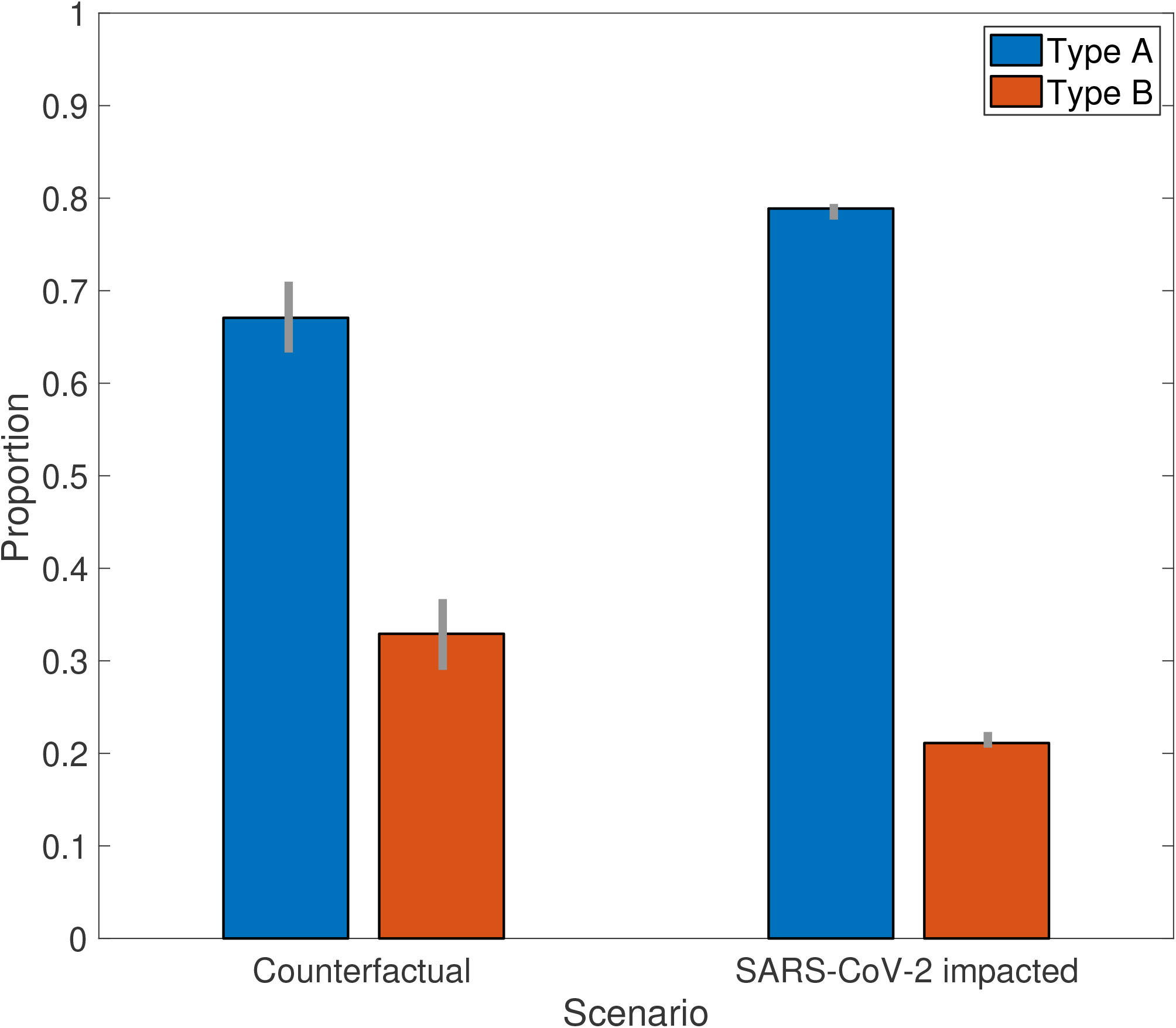
Estimated proportion of influenza cases in the overall population, during the 2021/2022 influenza season, attributed to type A influenza and type B influenza. Bars correspond to median estimates. Grey vertical bars correspond to 95% prediction intervals.

## 4 Discussion

The emergence of SARS-CoV-2 was followed by public health restrictions to inhibit the spread and limit the quantity of severe outcomes from COVID-19 disease. With possible knock-on effects of these interventions on the transmission of other respiratory viral infections, the situation posed questions as to the effects on upcoming influenza seasons. We have outlined our use of a previously developed seasonal influenza transmission model, calibrated to historical influenza seasons in England [10], to quantify the epidemiological implications on seasonal influenza (in England) of modified social interaction patterns and a broadened vaccination programme. We conducted these analyses prior to the 2020/2021 and 2021/2022 influenza seasons, respectively, contributing to the evidence base being compiled by scientific advisory groups in the UK that were aiding the COVID-19 pandemic response.

We forecasted that the combined interaction of NPIs (targeted at reducing COVID-19 transmission but also impacting influenza transmission) and a wide-spread and intensive vaccination campaign was likely to substantially reduce the scale of seasonal influenza epidemics over the winter - not only in terms of raw cases, but also in terms of hospitalisations and mortality. The combined action of NPIs and extended vaccination we anticipated may reduce influenza by over 95%. For the lead in to the 2020/2021 influenza season, with the intended coverage of the vaccination programme and interventions in place at the time the modelling suggesting that seasonal influenza was likely to pose a low burden to the health services was then borne out in England [8] and other regions in the temperate Northern Hemisphere [13, 14]; in the 2020/2021 influenza season the European Centre for Disease Prevention and Control stated that there were no hospitalised cases of influenza or fatalities from influenza reported by EU (European Union)/EEA (European Economic Area) countries [13], whilst in the USA the CDC reported that the cumulative rate of laboratory-confirmed influenza-associated hospitalisations was the lowest recorded since that type of data collection began [14].

Although a dramatic drop in harms from influenza was predicted in all ages, those aged 2-17 were predicted to experience the lowest decline. Consequently, it was suggested that any spare vaccine capacity (after vaccinating the eldest and those at-risk of complications if infected) should be targeted towards school-aged children; in the context of England, supplementing the current primary-school programme with vaccination in secondary schools.

Our result suggesting a compensatory influenza season in 2021/2022 due to a suppressed influenza season in 2020/2021 has also been demonstrated in other mathematical model frameworks. One such example was by Krauland *et al*. [15], who used an agent-based model and also investigated the relationship between the potential magnitude of a resurgent influenza epidemic to cross-immunity from past infection and transmissibility of strains. In another example, applied to the United States, Lee *et al*. [16] used a more parsimonious, non-age structured compartmental influenza transmission model that included immunity propagation. They also showed that higher influenza vaccine uptake would reduce the projected resurgence in influenza.

Public health interventions against SARS-CoV-2 have had a profound impact on the worldwide circulation of other respiratory viruses, including seasonal influenza [17]. As a result of non-pharmaceutical interventions in place for COVID-19 (such as mask-wearing, reduced social interactions and international travel) influenza activity levels were extremely low globally in 2020 to 2021 [18]. In the temperate Northern Hemisphere, during the warmer summer months of 2022 when rates of respiratory viral infections would normally be very low or absent, some countries saw unusual seasonal changes in the prevalence of influenza [19], respiratory syncytial virus (RSV) [20] and parainfluenza [21]; these observations coincided with a reduction in COVID-19 mitigation measures. In England, there has been indications of an early onset of the 2022/2023 influenza season, with the hospital admission rate for confirmed cases of influenza in mid-October being greater in 2022 than seen in preceding years [22].

Looking ahead, there is uncertainty as to the size of future seasonal influenza epidemics. The potential elimination of the B/Yamagata lineage [23] may have as yet unforeseen ramifications on the evolution of influenza strains and the multi-strain transmission dynamics. A multi-nation analysis by Ali *et al*. [24], using a data-driven mechanistic predictive modelling framework, suggested the potential for substantial increases in infection burden in upcoming influenza seasons worldwide. Meanwhile, results by Felix Garza *et al*. [25] suggest that the risks posed by seasonal influenza viruses remained largely unchanged during the first two years of the COVID-19 pandemic and that the sizes of future seasonal influenza virus epidemics will likely be similar to those observed before the pandemic.

Another area of doubt is the robustness of pathogen surveillance and data streams in perpetuity. The COVID-19 pandemic has impacted surveillance systems [26–28] and existing vaccination programmes [29–31], leading to uncertainty in how both influenza and COVID-19 surveillance can be jointly and durably implemented over prolonged time horizons. We must understand and assess any changes that impact influenza surveillance in order to inform public health for future actions.

Our choice of model structure and parameterisation inherently has associated assumptions. Thus, it is important our findings are viewed with due consideration of the limitations. We summarise here four of particular note. First, our model takes a national perspective, but regional variations in adherence to NPIs and/or vaccine uptake may be crucial. For example, areas where uptake of the seasonal influenza vaccine in target groups has been historically below the national average might also have differing NPI usage and uptake of an expanded vaccination programme compared to the national average. Second, and as previously mentioned, the framework did not explicitly include healthcare workers or care-home residents and staff, who can be particularly at risk of being exposed to seasonal influenza. Third, in the first piece of analysis undertaken prior to the 2020/2021 influenza season, to include the impact of NPIs on the age-structure of contacts in the relevant scenarios, we used estimates that included schools being open, with this amended contact structure then being used throughout the duration of the influenza season. We did not incorporate temporally varying contact patterns to capture school holidays, or the possibility of social mixing patterns changing around dates tied to special occasions (the Christmas and New Year period, for example). Finally, given the relative novelty of the SARS-CoV-2 virus in humans, there remains a great amount of uncertainty in our understanding of the interaction of SARS-CoV-2 with other respiratory infections in circulation, such as influenza [32]. We did not consider the consequences of interaction between SARS-CoV-2 and seasonal influenza, such as coinfections. Our knowledge on processes such as viral interference and coinfection will continue to improve as new information is accumulated.

It is not yet apparent when a more predictable pattern of respiratory virus circulation, including seasonal influenza, will return; over time we will develop an improved understanding of how circulatory patterns of respiratory viruses (and other pathogens) have altered in the context of the COVID-19 pandemic. Ongoing consideration of preventive measures to reduce influenza virus infection risks in the community is therefore prudent, including strengthening influenza vaccination programmes. Scenario assessments using epidemiological models and mathematical models of infectious disease dynamics can continue to provide quantitative evidence for the potential impact of different epidemiological characteristics and public health policies on the course of seasonal influenza outbreaks, particularly following receivership of new epidemiological and behavioural data.

## Data Availability

All data utilised in this study are publicly available, with relevant references and data repositories stated within the main manuscript and Supporting Information.
The code repository for the study is available on GitHub: https://github.com/WarwickSBIDER/diminished-influenza-seasons.

## Author contributions

**Edward M. Hill:** Conceptualisation, Data curation, Formal analysis, Methodology, Software, Validation, Visualisation, Writing - Original Draft, Writing - Review &Editing.

**Matt J. Keeling:** Funding acquisition, Methodology, Supervision, Visualisation, Writing - Review &Editing.

## Financial disclosure

EMH and MJK were supported by the Medical Research Council through the COVID-19 Rapid Response Rolling Call [grant number MR/V009761/1] and the Biotechnology and Biological Sciences Research Council [grant number: BB/S01750X/1]. MJK was supported by the Vaccine Efficacy Evaluation for Priority Emerging Diseases (VEEPED) project through the National Institute for Health Research using Official Development Assistance (ODA) funding. MJK was supported by UKRI through the JUNIPER modelling consortium [grant number MR/V038613/1] and by the Engineering and Physical Sciences Research Council through the MathSys CDT [grant number EP/S022244/1]. MJK was also supported by the National Institute for Health Research (NIHR) [Policy Research Programme, Mathematical &Economic Modelling for Vaccination and Immunisation Evaluation, and Emergency Response; NIHR200411]. The views expressed are those of the authors and not necessarily those of the NIHR or the Department of Health and Social Care. MJK is affiliated to the National Institute for Health Research Health Protection Research Unit (NIHR HPRU) in Gastrointestinal Infections at University of Liverpool in partnership with UK Health Security Agency (UKHSA), in collaboration with University of Warwick. MJK is also affiliated to the National Institute for Health Research Health Protection Research Unit (NIHR HPRU) in Genomics and Enabling Data at University of Warwick in partnership with UK Health Security Agency (UKHSA). The views expressed are those of the author(s) and not necessarily those of the NHS, the NIHR, the Department of Health and Social Care or UK Health Security Agency. The funders had no role in study design, data collection and analysis, decision to publish, or preparation of the manuscript.

## Data availability

All data utilised in this study are publicly available, with relevant references and data repositories stated within the main manuscript and Supporting Information.

## Code availability

The code repository for the study is available on GitHub: https://github.com/WarwickSBIDER/diminished-influenza-seasons.

## Competing interests

All authors declare that they have no competing interests.

## Notes

### Competing Interest Statement

The authors have declared no competing interest.

### Funding Statement

EMH and MJK were supported by the Medical Research Council through the COVID-19 Rapid Response Rolling Call [grant number MR/V009761/1] and the Biotechnology and Biological Sciences Research Council [grant number: BB/S01750X/1]. MJK was supported by the Vaccine Efficacy Evaluation for Priority Emerging Diseases (VEEPED) project through the National Institute for Health Research using Official Development Assistance (ODA) funding. MJK was supported by UKRI through the JUNIPER modelling consortium [grant number MR/V038613/1] and by the Engineering and Physical Sciences Research Council through the MathSys CDT [grant number EP/S022244/1]. MJK was also supported by the National Institute for Health Research (NIHR) [Policy Research Programme, Mathematical \& Economic Modelling for Vaccination and Immunisation Evaluation, and Emergency Response; NIHR200411]. The views expressed are those of the authors and not necessarily those of the NIHR or the Department of Health and Social Care. MJK is affiliated to the National Institute for Health Research Health Protection Research Unit (NIHR HPRU) in Gastrointestinal Infections at University of Liverpool in partnership with UK Health Security Agency (UKHSA), in collaboration with University of Warwick. MJK is also affiliated to the National Institute for Health Research Health Protection Research Unit (NIHR HPRU) in Genomics and Enabling Data at University of Warwick in partnership with UK Health Security Agency (UKHSA). The views expressed are those of the author(s) and not necessarily those of the NHS, the NIHR, the Department of Health and Social Care or UK Health Security Agency. The funders had no role in study design, data collection and analysis, decision to publish, or preparation of the manuscript.

## References

[1] Bellizzi S, Panu Napodano CM, Pinto S, Pichierri G. COVID-19 and seasonal influenza: The potential 2021–22 “Twindemic”. Vaccine 40(24):3286–3287 (2022). doi:10.1016/j.vaccine.2022.04.074.

[2] Cowling BJ, Ali ST, Ng TWY, Tsang TK, Li JCM, et al. Impact assessment of non-pharmaceutical interventions against coronavirus disease 2019 and influenza in Hong Kong: an observational study. The Lancet Public Health 5(5):e279–e288 (2020). doi:10.1016/S2468-2667(20)30090-6.

[3] Emborg HD, Carnahan A, Bragstad K, Trebbien R, Brytting M, et al. Abrupt termination of the 2019/20 influenza season following preventive measures against COVID-19 in Denmark, Norway and Sweden. Eurosurveillance 26(22):pii=2001160 (2021). doi:10.2807/1560-7917.ES.2021.26.22.2001160.

[4] Sullivan SG, Carlson S, Cheng AC, Chilver MB, Dwyer DE, et al. Where has all the influenza gone? The impact of COVID-19 on the circulation of influenza and other respiratory viruses, Australia, March to September 2020. Eurosurveillance 25(47):pii=2001847 (2020). doi:10.2807/1560-7917.ES.2020.25.47.2001847.

[5] World Health Organization. Influenza Update N° 375 (2020). URL https://cdn.who.int/media/docs/default-source/influenza/influenza-updates/2020/20200831surveillanceupdate375.pdf?sfvrsn=3df6955a7. [Online] (Accessed: 13 October 2022).

[6] McCauley J, Barr IG, Nolan T, Tsai T, Rockman S, et al. The importance of influenza vaccination during the COVID-19 pandemic. Influenza and Other Respiratory Viruses 16(1):3–6 (2022). doi: 10.1111/irv.12917.

[7] Department of Health and Social Care. National flu immunisation programme 2020 to 2021 letter: update (2020). URL https://webarchive.nationalarchives.gov.uk/ukgwa/20210701160234/ https://www.gov.uk/government/publications/national-flu-immunisation-programme-plan. [Online] (Accessed: 13 October 2022).

[8] Public Health England. Surveillance of influenza and other seasonal respiratory viruses in the UK: Winter 2020 to 2021 (2021). URL https://webarchive.nationalarchives.gov.uk/ukgwa/20220401215804/ https://www.gov.uk/government/statistics/annual-flu-reports. [Online] (Accessed: 13 October 2022).

[9] Welsh Government Technical Advisory Cell. Technical Advisory Group: winter modelling update 10 September 2021 (2021). URL https://gov.wales/sites/default/files/publications/2021-09/technical-advisory-group-winter-modelling-update-10-september-2021.pdf. [Online] (Accessed: 13 October 2022).

[10] Hill EM, Petrou S, Forster H, de Lusignan S, Yonova I, et al. Optimising age coverage of seasonal influenza vaccination in England: A mathematical and health economic evaluation. PLOS Computational Biology 16(10):e1008278 (2020). doi:10.1371/journal.pcbi.1008278.

[11] Keeling MJ, Dyson L, Guyver-Fletcher G, Holmes A, Semple MG, et al. Fitting to the UK COVID-19 outbreak, short-term forecasts and estimating the reproductive number. Statistical Methods in Medical Research 31(9):1716–1737 (2022). doi:10.1177/09622802211070257.

[12] Department of Health and Social Care. National flu immunisation programme 2021 to 2022 letter (2021). URL https://webarchive.nationalarchives.gov.uk/ukgwa/20220412180617/ https://www.gov.uk/government/publications/national-flu-immunisation-programme-plan/national-flu-immunisation-programme-2021-to-2022-letter. [Online] (Accessed: 13 October 2022).

[13] European Centre for Disease Prevention and Control. Seasonal influenza - Annual Epidemiological Report for 2020-2021 (2021). URL https://www.ecdc.europa.eu/sites/default/files/documents/AER-seasonal-influenza-2020-final.pdf. [Online] (Accessed: 13 October 2022).

[14] Centres for Disease Control and Prevention. 2020-2021 Flu Season Summary (2021). URL https://www.cdc.gov/flu/season/faq-flu-season-2020-2021.htm. [Online] (Accessed: 13 October 2022).

[15] Krauland MG, Galloway DD, Raviotta JM, Zimmerman RK, Roberts MS. Impact of Low Rates of Influenza on Next-Season Influenza Infections. American Journal of Preventive Medicine 62(4):503–510 (2022). doi:10.1016/j.amepre.2021.11.007.

[16] Lee K, Jalal H, Raviotta JM, Krauland MG, Zimmerman RK, et al. Estimating the Impact of Low Influenza Activity in 2020 on Population Immunity and Future Influenza Seasons in the United States. Open Forum Infectious Diseases 9(1):ofab607 (2022). doi:10.1093/ofid/ofab607.

[17] The Lancet Respiratory Medicine. COVID-19 pandemic disturbs respiratory virus dynamics. The Lancet Respiratory Medicine 10(8):725 (2022). doi:10.1016/S2213-2600(22)00255-7.

[18] Bonacina F, Bo&eumllle PY, Colizza V, Lopez O, Thomas M, et al. Global patterns and drivers of influenza decline during the COVID-19 pandemic. medRxiv page 2022.07.15.22277497 (2022). doi:10.1101/2022.07.15.22277497.

[19] World Health Organization. Influenza Update N° 422 (2022). URL https://cdn.who.int/media/docs/default-source/influenza/influenza-updates/2022/20220627surveillanceupdate422.pdf?sfvrsn=e14b0141. [Online] (Accessed: 13 October 2022).

[20] Eden JS, Sikazwe C, Xie R, Deng YM, Sullivan SG, et al. Off-season RSV epidemics in Australia after easing of COVID-19 restrictions. Nature Communications 13:2884 (2022). doi:10.1038/s41467-022-30485-3.

[21] Kuitunen I, Artama M, Haapanen M, Renko M. Record high parainfluenza season in children after relaxation of COVID-19 restrictions in fall 2021 - A nationwide register study in Finland. Influenza and Other Respiratory Viruses 16(4):613–616 (2022). doi:10.1111/irv.12983.

[22] UK Health Security Agency. Weekly national Influenza and COVID-19 surveillance report: Week 42 report (up to week 41 data) 20 October 2022 (2022). URL https://assets.publishing.service.gov.uk/government/uploads/system/uploads/attachmentdata/file/1112388/WeeklyFluandCOVID-19reportw42.pdf. [Online] (Accessed: 25 October 2022).

[23] Dhanasekaran V, Sullivan S, Edwards KM, Xie R, Khvorov A, et al. Human seasonal influenza under COVID-19 and the potential consequences of influenza lineage elimination. Nature Communications 13:1721 (2022). doi:10.1038/s41467-022-29402-5.

[24] Ali ST, Lau YC, Shan S, Ryu S, Du Z, et al. Prediction of upcoming global infection burden of influenza seasons after relaxation of public health and social measures during the COVID-19 pandemic: a modelling study. The Lancet Global Health 10(11):e1612–e1622 (2022). doi: 10.1016/S2214-109X(22)00358-8.

[25] Felix Garza ZC, de Jong SPJ, Gibson J, Han AX, van Leeuwen S, et al. Impacts of the COVID-19 pandemic on future seasonal influenza epidemic. medRxiv page 2022.02.05.22270494 (2022). doi:10.1101/2022.02.05.22270494.

[26] Adlhoch C, Sneiderman M, Martinuka O, Melidou A, Bundle N, et al. Spotlight influenza: The 2019/20 influenza season and the impact of COVID-19 on influenza surveillance in the WHO European Region. Eurosurveillance 26(40):pii=2100077 (2021). doi:https://doi.org/10.2807/1560-7917.ES.2021.26.40.2100077.

[27] Rigoine de Fougerolles T, Puig-Barbera J, Kassianos G, Vanhems P, Schelling J, et al. A comparison of coronavirus disease 2019 and seasonal influenza surveillance in five European countries: France, Germany, Italy, Spain and the United Kingdom. Influenza and Other Respiratory Viruses 16(3):417–428 (2022). doi:10.1111/irv.12941.

[28] Sabeena S, Ravishankar N, Robin S. The impact of COVID-19 pandemic on influenza surveillance: a systematic review and meta-analysis. medRxiv page 2022.03.31.22273236 (2022). doi:10.1101/2022.03.31.22273236.

[29] Shet A, Carr K, Danovaro-Holliday MC, Sodha SV, Prosperi C, et al. Impact of the SARS-CoV-2 pandemic on routine immunisation services: evidence of disruption and recovery from 170 countries and territories. The Lancet Global Health 10(2):e186–e194 (2022). doi:10.1016/S2214-109X(21)00512-X.

[30] Causey K, Fullman N, Sorensen RJD, Galles NC, Zheng P, et al. Estimating global and regional disruptions to routine childhood vaccine coverage during the COVID-19 pandemic in 2020: a modelling study. The Lancet 398(10299):522–534 (2021). doi:10.1016/S0140-6736(21)01337-4.

[31] Gaythorpe KA, Abbas K, Huber J, Karachaliou A, Thakkar N, et al. Impact of COVID-19-related disruptions to measles, meningococcal A, and yellow fever vaccination in 10 countries. eLife 10(10299):469–471 (2021). doi:10.7554/eLife.67023.

[32] Heinzinger S, Eberle U, Angermeier H, Flechsler J, Konrad R, et al. Reciprocal circulation pattern of SARS-CoV-2 and influenza viruses during the influenza seasons 2019/2020 and 2020/2021 in the Bavarian Influenza Sentinel (Germany). Epidemiology and Infection 149:e226 (2021). doi: 10.1017/S0950268821002296.

