## Supporting Information for "Scenario modelling for diminished influenza seasons during 2020/2021 and 2021/2022 in England"

**Additional figures** **7**

**Additional tables** **10**

#### Complementary details of the epidemiological modelling approach

This is verbatim from the Supporting Information of Hill *et al.* [1].

##### Model framework schematic

We provide below a visual depiction of the entire model structure and incorporation of the data streams within and between the four model components: vaccination model, immunity propagation model, epidemiological model and observation model (Fig. S1).

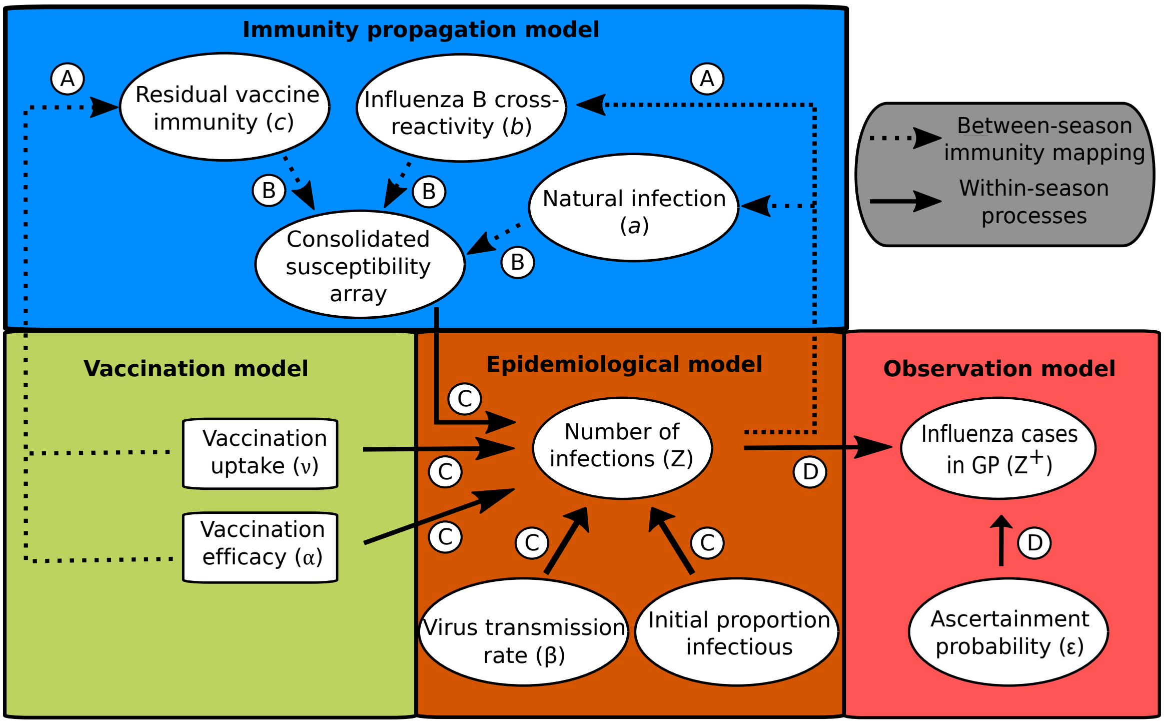

**Fig. S1: Schematic showing the links between the vaccination, immunity propagation, epidemiological and observation model components.** We adopt the visualisation conventions of [2], with ellipses indicating variables, and rectangles indicating data. Dotted arrows indicate relationships between prior season epidemiological outcomes and immunity propagation factors. Solid arrows indicate within-season processes. Circled capitalised letters indicate the relationships connecting the variables or data involved. These relationships are: process A, propagation of immunity as a result of exposure to influenza virus in the previous influenza season (through natural infection or vaccination); process B, modulation of current influenza season virus susceptibility; process C, estimation of influenza case load via the SEIR model of transmission; process D, ascertainment of cases through ILI recording at GP. Reproduced from Hill *et al.* [3].

##### Immunity propagation model

We had a total of ten exposure history groupings and associated strain-specific susceptibilities, which we consolidated into a single susceptibility array (Fig. S2).

We expand here our protocol for parameterisation of susceptibility values amongst exposure groups capturing those both vaccinated and naturally infected in the prior season (Fig. S2: rows 7–10).

For this collection of exposure histories, susceptibility to a subset of strains may conceivably be collectively modified via natural infection and vaccination immunity propagation pathways. In these instances, we treated the immunity propagation mechanisms independently, with the modified susceptibility set by the dominant immunity propagation entity (i.e. $\min(a, c_m)$, $\min(b, c_m)$). In other

| | | Strain susceptibility | | | |
|---|---|---|---|---|---|
| | | A(H1N1)pdm09 | A(H3N2) | B/Victoria | B/Yamagata |
| Exposure history (h) | Naïve | 1 | 1 | 1 | 1 |
| | A(H1N1)pdm09 | $a$ | 1 | 1 | 1 |
| | A(H3N2) | 1 | $a$ | 1 | 1 |
| | B/Victoria | 1 | 1 | $a$ | $b$ |
| | B/Yamagata | 1 | 1 | $b$ | $a$ |
| | Vacc. (V) | $c_{\mathrm{A(H1N1)}}$ | $c_{\mathrm{A(H3N2)}}$ | $c_{\mathrm{B/Victoria}}$ | $c_{\mathrm{B/Yamagata}}$ |
| | A(H1N1)pdm09 & V | $\min(a, c_{\mathrm{A(H1N1)}})$ | $c_{\mathrm{A(H3N2)}}$ | $c_{\mathrm{B/Victoria}}$ | $c_{\mathrm{B/Yamagata}}$ |
| | A(H3N2) & V | $c_{\mathrm{A(H1N1)}}$ | $\min(a, c_{\mathrm{A(H3N2)}})$ | $c_{\mathrm{B/Victoria}}$ | $c_{\mathrm{B/Yamagata}}$ |
| | B/Victoria & V | $c_{\mathrm{A(H1N1)}}$ | $c_{\mathrm{A(H3N2)}}$ | $\min(a, c_{\mathrm{B/Victoria}})$ | $\min(b, c_{\mathrm{B/Yamagata}})$ |
| | B/Yamagata & V | $c_{\mathrm{A(H1N1)}}$ | $c_{\mathrm{A(H3N2)}}$ | $\min(b, c_{\mathrm{B/Victoria}})$ | $\min(a, c_{\mathrm{B/Yamagata}})$ |

**Fig. S2: Infographic presenting the interaction between exposure history and susceptibility.** The interaction between exposure history $h$ and susceptibility to strain $m$ in the current season, $f(h, m)$, was classified into ten distinct groups: One group for the naive (uninfected and not vaccinated, row 1); one group per strain, infected but not vaccinated (rows 2–5); one group for those vaccinated and experiencing no natural infection (row 6); one group per strain for being infected and vaccinated (rows 7–10). We let $a$ denote modified susceptibility to strain $m$ given infection by a strain $m$ type virus the previous season (dark green shading), $b$ modified susceptibility due to cross-reactivity between type B influenza lineages (dark blue shading), and $c_m$ the change in susceptibility to strain $m$ given vaccination in the previous season (gold shading). Unmodified susceptibilities retained a value of 1 (red shading). We enforced $0 < a,b,c_m < 1$. Reproduced from Hill *et al.* [3].

words, we took a pessimistic stance by assuming no boosting of the immunity propagation response as a result of dual influenza virus exposure (from both natural infection and vaccination).

#### Epidemiological model specifics

The epidemiological model was a deterministic, age-dependent (single year age brackets), multi-strain structured compartmental based model capturing influenza infection status (with susceptible-latent-infected-recovered, SEIR, dynamics) and vaccine uptake. In addition, we assumed disease transmission to be frequency-dependent.

We let $E_i^{X,m}$, $I_i^{X,m}$ and $R_i^{X,m}$ denote the proportion of the population that were in age class $i$, with vaccination status $X \in \{N, V\}$, and that were latent, infectious and recovered (as a result of natural infection) with respect to strain $m$.

Exposure to influenza virus in the previous influenza season, through natural infection or vaccination, modulated current influenza season susceptibility. Tracking immunity derived from natural infection and vaccination separately required, per age bracket, ten distinct exposure history groups, with the susceptibility to strain $m$ for a given exposure history group $h$ encoded into the susceptibility array $f(h, m)$.

To incorporate exposure history from the previous influenza season, we let $S_i^{X,h}$ denote the proportion of the population that are in age class $i$, with exposure history $h$ and vaccination status $X \in \{N, V\}$ that are susceptible to all strains. With ten exposure history groups $h$ in place, a total of 20 susceptibility states were used: ten $S^{N,h}$ states, accounting for susceptibles not vaccinated in the current influenza season stratified by exposure history grouping; ten $S^{V,h}$ states, tracking susceptibles who had been administered the vaccine in the current influenza season whilst retaining exposure history group information.

The epidemiological model had the following formulation, with time dependencies dropped:

$$
\begin{cases}
\frac{dS_i^N}{dt} & = -\left(\sum_m \left(\sum_h f(h,m) S_i^{N,h}\right) \lambda_i^m\right) - \mu_i S_i^N \\
\frac{dS_i^V}{dt} & = -\left(\sum_m \left(\sum_h f(h,m) S_i^{V,h}\right) (1-\alpha_i^m)\lambda_i^m\right) + \mu_i S_i^N \\
\frac{dE_i^{N,m}}{dt} & = \left(\sum_h f(h,m) S_i^{N,h}\right) \lambda_i^m - \gamma_1^m E_i^{N,m} - \mu_i E_i^{N,m} \\
\frac{dI_i^{N,m}}{dt} & = \gamma_1^m E_i^{N,m} - \gamma_2 I_i^{N,m} - \mu_i I_i^{N,m} \\
\frac{dR_i^{N,m}}{dt} & = \gamma_2 I_i^{N,m} - \mu_i R_i^{N,m} \\
\frac{dE_i^{V,m}}{dt} & = \left(\sum_h f(h,m) S_i^{V,h}\right) (1-\alpha_i^m)\lambda_i^m - \gamma_1^m E_i^{V,m} + \mu_i E_i^{N,m} \\
\frac{dI_i^{V,m}}{dt} & = \gamma_1^m E_i^{V,m} - \gamma_2 I_i^{V,m} + \mu_i I_i^{N,m} \\
\frac{dR_i^{V,m}}{dt} & = \gamma_2 I_i^{V,m} + \mu_i R_i^{N,m}
\end{cases}
\tag{1}
$$

where $\gamma_1^m$ corresponds to the rate of loss of latency (strain dependent), $\gamma_2$ the rate of loss of infectiousness (strain independent), $\alpha_i^m$ the age-specific vaccine efficacy for strain $m$, and $\mu_i = \frac{\nu_i}{S_i^N + \sum_m (E_i^{N,m} + I_i^{N,m} + R_i^{N,m})}$ with $\nu_i$ the rate of immunisation in age group $i$.

We expressed the age and strain specific force of infection for age group $i$ and strain $m$, $\lambda_i^m$, as follows,

$$
\lambda_i^m = q_m \sigma_i^m \sum_j \sum_X c_{ij} \frac{I_j^{X,m}}{n_j},
$$

where $q_m$ represents the transmissibility parameter for strain $m$ (imputed from the contact structure, recovery rate, and the strain-specific basic reproduction number), $\sigma_i^m$ the susceptibility of age group $i$ towards strain $m$, $c_{ij}$ the frequency at which individuals in age group $i$ make contact with those in age group $j$ (see Section 2.3.1 for details on parameterisation), and $n_j$ the proportion of the population in age group $j$.

##### Contact matrix

The structure of social contacts were taken from Fumanelli *et al.* [4], who simulated a population of synthetic individuals in order to derive frequencies of total contacts by age (in other words, the total number of contacts between individuals of age $i$ and individuals of age $j$) for 26 European countries (including the United Kingdom).

Ultimately, we sought estimates of the average number of contacts a *given* person of age $i$ had with people of aged $j$. Thus, we transformed the published symmetric matrix of adequate contacts in the United Kingdom ([4]) by dividing the matrix by the age structure of the population; the result was a contact matrix $c_{i,j}$ providing the average number of adequate contacts an individual of age group $i$ has with individuals of age group $j$.

The United Kingdom data was stratified into yearly age groups up to the single year age bracket 89–90. With our age-structured transmission model including ages 90–100+, the following assumptions were enforced: the average number of adequate contacts individuals residing in ages 90–100+ had with individuals of age group $j$ were set equivalent to the adequate contact quantities listed for the

89–90 age bracket ($c_{91:101,1:90} = c_{90,1:90}$); the average number of adequate contacts individuals residing in ages 0-90 had with individuals aged 90+ equalled the average number of adequate contacts had with those aged 89–90 $c_{1:90,91:101} = c_{1:90,90}$; all single-year age bracket interactions for those aged 90+ took the value of average number of adequate contacts an individual aged 89–90 had with people aged 89–90 ($c_{91:101,91:101} = c_{90,90}$).

**Between-season exposure history group mappings**

On 1st September we perform the following updates (for clarity, the dependencies on age-group have been dropped):

- $S^N \to S^{N,h=\bar{N}}$
- $S^V \to S^{N,h=\bar{V}}$
- $\{E^{N,m}, I^{N,m}, R^{N,m}\} \to S^{N,h=m}$
- $\{E^{V,m}, I^{V,m}, R^{V,m}\} \to S^{N,h=m/\bar{V}}$

where $X$ represents vaccination status, with $N$ corresponding to naive/non-vaccinated and $V$ vaccinated.

**End of influenza season demographic processes**

Although the epidemiological model ODEs were devoid of explicit demographic processes, we instead implemented fluctuations to the age structure (due to demographic processes) prior to the initiation of each influenza season.

For the time period 2010 to 2018 inclusive, we informed age-stratified population distributions for each influenza season from annual mid-year ONS population estimates [5]. Revised values for the proportion of the population contained within each unvaccinated susceptible class ($S^{N,h}$), at the outset of the influenza season, could subsequently be computed. We scaled prior influenza season values up or down as required to ensure the updated age-level proportion was matched. In addition, we assumed that, per age bracket, overall increases or decreases in population size were apportioned amongst the exposure history entities weighted according to the end-of-season exposure history proportions.

Mathematically, the entity $S_i^{N,h}$ obeyed the following,

$$S_i^{N,h} = \hat{S}_i^{N,h} \times \text{scale}_i(t+1) \quad \forall\, i \in \{1, 2, \ldots, 99, 100\}$$

where $\hat{S}_i^{N,h}$ corresponded to the start of influenza season unvaccinated susceptible class value for age class $i$ and exposure history group $h$ before population adjustments (i.e. demographic processes) were applied, and

$$\text{scale}_i(t+1) = \begin{cases} n_i(t+1)/n_{i-1}(t), & \text{if } i \in \{1, 2, \ldots, 98, 99\}. \\ n_i(t+1)/(n_{i-1}(t) + n_i(t)), & \text{if } i = 100. \end{cases} \tag{2}$$

### Case severity assumptions

We summarise here the values applied for mapping infections to three different levels of case severity: (i) case ascertainment; (ii) hospitalisation (inpatient admissions); (iii) mortality. A description of the sources used for the parameterisation of the case severity module may be found in Hill *et al.* [1].

#### Case ascertainment

To translate infections to a measure indicating the amount of people who had severe enough symptoms to consult a GP, we used an age-dependent ascertainment probability. The age-dependent ascertainment probabilities were taken relative to those aged 100+yrs, who took value 1. We then applied a piecewise linear profile to acquire an age-dependent ascertainment curve.

Notch ages were 0, 2, 18, 65, 85. Median values at the notch ages were [0.2087, 0.1383, 0.2367, 0.6974, 0.5116].

For those categorised as at-risk, we applied a scale factor of 1.5 to the ascertainment probability.

#### Hospital inpatient admissions

To the ascertained cases, we applied a scaling to inform the relative number of inpatient hospital admissions attributable to influenza. These estimates were influenza type, risk group and age specific (Table S1).

**Table S1: Inpatient admission event occurrence relative to influenza attributed GP visits.** Estimates are influenza type, risk group and age group specific. All values are displayed to 2 decimal places.

| | **Type A** | | **Type B** | |
|---|---|---|---|---|
| **Age (yrs)** | **low-risk** | **at-risk** | **low-risk** | **at-risk** |
| **0-1** | 0.79 | 4.16 | 0.81 | 3.46 |
| **2-5** | 0.13 | 0.88 | 0.13 | 0.85 |
| **6-9** | 0.06 | 0.34 | 0.05 | 0.32 |
| **10-19** | 0.06 | 0.18 | 0.00 | 0.00 |
| **20-29** | 0.00 | 0.19 | 0.00 | 0.00 |
| **30-39** | 0.07 | 0.22 | 0.00 | 0.00 |
| **40-49** | 0.06 | 0.27 | 0.00 | 0.00 |
| **50-59** | 0.07 | 0.33 | 0.00 | 0.00 |
| **60-64** | 0.06 | 0.36 | 0.00 | 0.00 |
| **65-74** | 0.11 | 0.74 | 0.00 | 0.00 |
| **75-84** | 0.31 | 1.01 | 0.00 | 0.00 |
| **85+** | 1.27 | 1.47 | 0.00 | 0.00 |

#### Mortality

*In hospital mortality*

To the ascertained cases, we applied a scaling to inform the relative number of in hospital deaths from influenza. These estimates were influenza type, risk group and age specific (Table S2).

**Table S2: In hospital mortality event occurrence relative to influenza attributed GP visits.** Estimates are influenza type, risk group and age group specific. We assumed no deaths were caused by influenza B. All values are displayed to 4 decimal places.

| | **Type A** | | **Type B** | |
|---|---|---|---|---|
| **Age (yrs)** | **low-risk** | **at-risk** | **low-risk** | **at-risk** |
| **0-1** | 0.0020 | 0.2025 | 0.0000 | 0.0000 |
| **2-5** | 0.0000 | 0.0300 | 0.0000 | 0.0000 |
| **6-9** | 0.0000 | 0.0090 | 0.0000 | 0.0000 |
| **10-19** | 0.0002 | 0.0040 | 0.0000 | 0.0000 |
| **20-29** | 0.0001 | 0.0030 | 0.0000 | 0.0000 |
| **30-39** | 0.0001 | 0.0060 | 0.0000 | 0.0000 |
| **40-49** | 0.0005 | 0.0200 | 0.0000 | 0.0000 |
| **50-59** | 0.0005 | 0.0200 | 0.0000 | 0.0000 |
| **60-64** | 0.0010 | 0.0300 | 0.0000 | 0.0000 |
| **65-74** | 0.0020 | 0.0800 | 0.0000 | 0.0000 |
| **75-84** | 0.0130 | 0.1400 | 0.0000 | 0.0000 |
| **85+** | 0.1310 | 0.3300 | 0.0000 | 0.0000 |

*Out of hospital mortality*

We applied the following age-dependent ratios of mortality events occurring out of hospital compared to in-hospital:

- Ages 0-49: 100% in hospital, 0% out of hospital;
- Ages 50-64: 75% in hospital, 25% out of hospital;
- Ages 65-74: 65% in hospital, 35% out of hospital;
- Ages 75+: 50% in hospital, 50% out of hospital.

### Additional figures

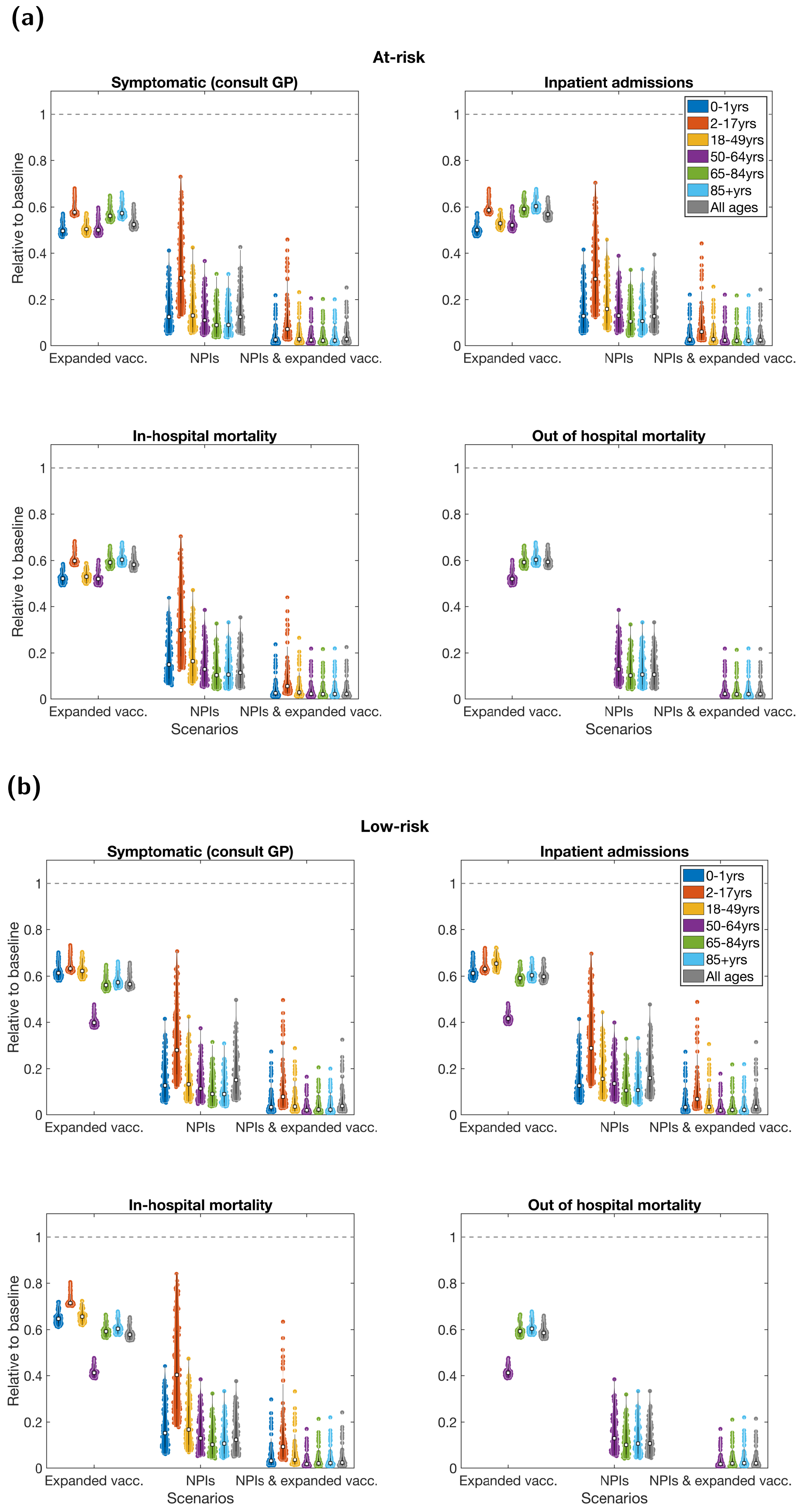

**Fig. S3: For the 2020/2021 influenza season scenarios, age- and risk-stratified health episode occurrences relative to the baseline scenario.** The displayed panels consider the fraction of the population that were: **(a)** at-risk; **(b)** low-risk. Each age grouping is measured relative to the outcomes attained for that age grouping under the counterfactual scenario. In each panel, we batch the violin plots into three groups, corresponding to one of the three non-baseline scenarios: the left group for the expanded vaccination scenario, the central group for the NPIs scenario and the right group for the combined NPIs and expanded vaccination scenario. White squares represent the medians. Solid black lines the interquartile range. See Table S3 for median values and 95% prediction intervals.

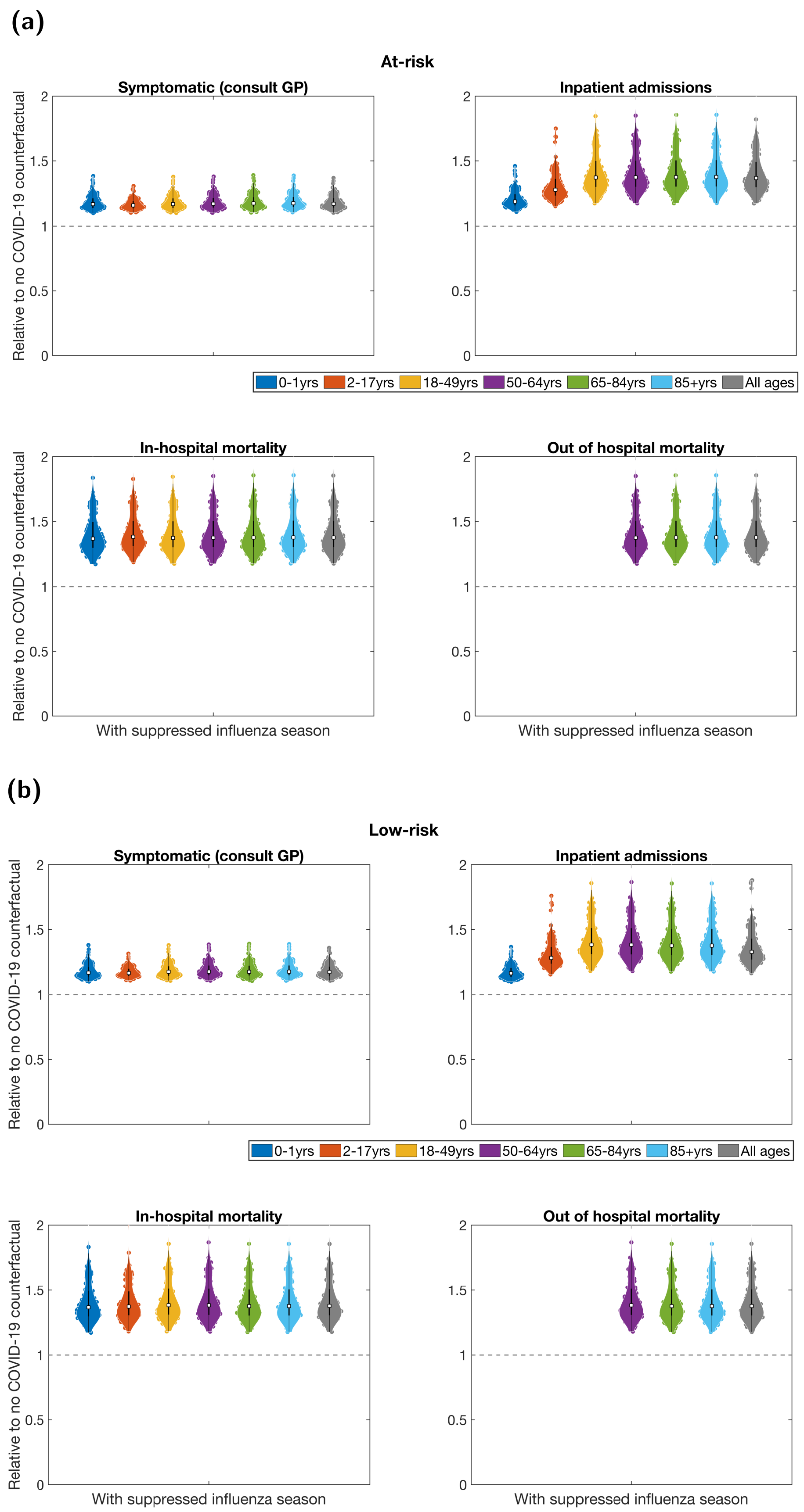

**Fig. S4: For the 2021/2022 influenza season scenarios, age- and risk-stratified health episode occurrences relative to the non-COVID counterfactual scenario.** The displayed panels consider the fraction of the population that were: **(a)** at-risk; **(b)** low-risk. Each age grouping is measured relative to the outcomes attained for that age grouping under the counterfactual scenario. White squares represent the medians. Solid black lines the interquartile range. See Table S4 for median values and 95% prediction intervals.

### Additional tables

**Table S3:** For the pre-2020/2021 influenza season analysis, summary statistics for the age-stratified health episode occurrences as proportions relative to the baseline scenario. For the 100 simulations performed per scenario, we report the medians and in parentheses the 95% prediction intervals. We report estimates for each statistic to two decimal places. We assumed there were no out of hospital fatalities due to seasonal influenza for 0-49 year olds, with corresponding cells having value N/A.

| | | | Age group (years) | | | | | | |
|---|---|---|---|---|---|---|---|---|---|
| **Risk status** | **Statistic** | **Scenario** | **0-1** | **2-17** | **18-49** | **50-64** | **65-84** | **85+** | **All** |
| Overall population | Symptomatic (consult GP) | Expanded vaccination | 0.61 (0.58,0.69) | 0.62 (0.61,0.71) | 0.59 (0.56,0.67) | 0.44 (0.41,0.51) | 0.56 (0.54,0.64) | 0.57 (0.55,0.65) | 0.55 (0.53,0.63) |
| | | NPIs | 0.13 (0.06,0.34) | 0.28 (0.13,0.63) | 0.13 (0.06,0.34) | 0.11 (0.05,0.30) | 0.09 (0.04,0.24) | 0.09 (0.04,0.24) | 0.14 (0.06,0.38) |
| | | NPIs & expanded vaccination | 0.03 (0.01,0.20) | 0.08 (0.03,0.40) | 0.03 (0.01,0.20) | 0.02 (0.01,0.13) | 0.02 (0.01,0.14) | 0.02 (0.01,0.14) | 0.03 (0.01,0.22) |
| | Inpatient admissions | Expanded vaccination | 0.60 (0.57,0.68) | 0.61 (0.60,0.69) | 0.59 (0.56,0.65) | 0.50 (0.47,0.57) | 0.59 (0.56,0.66) | 0.60 (0.58,0.67) | 0.58 (0.55,0.65) |
| | | NPIs | 0.13 (0.06,0.34) | 0.29 (0.14,0.62) | 0.16 (0.07,0.37) | 0.13 (0.06,0.32) | 0.10 (0.05,0.26) | 0.11 (0.05,0.26) | 0.14 (0.06,0.34) |
| | | NPIs & expanded vaccination | 0.03 (0.01,0.20) | 0.06 (0.03,0.37) | 0.03 (0.01,0.20) | 0.02 (0.01,0.15) | 0.02 (0.01,0.15) | 0.02 (0.01,0.15) | 0.03 (0.01,0.19) |
| | In-hospital mortality | Expanded vaccination | 0.55 (0.52,0.62) | 0.61 (0.59,0.69) | 0.54 (0.51,0.60) | 0.52 (0.49,0.59) | 0.59 (0.56,0.66) | 0.60 (0.58,0.67) | 0.58 (0.56,0.65) |
| | | NPIs | 0.15 (0.07,0.37) | 0.31 (0.14,0.64) | 0.16 (0.07,0.39) | 0.13 (0.06,0.31) | 0.10 (0.05,0.26) | 0.11 (0.05,0.26) | 0.12 (0.05,0.29) |
| | | NPIs & expanded vaccination | 0.03 (0.01,0.18) | 0.06 (0.02,0.36) | 0.03 (0.01,0.19) | 0.02 (0.01,0.15) | 0.02 (0.01,0.15) | 0.02 (0.01,0.15) | 0.02 (0.01,0.15) |
| | Out of hospital mortality | Expanded vaccination | N/A | N/A | N/A | 0.52 (0.49,0.59) | 0.59 (0.57,0.66) | 0.60 (0.58,0.67) | 0.59 (0.57,0.66) |
| | | NPIs | N/A | N/A | N/A | 0.13 (0.06,0.31) | 0.10 (0.05,0.25) | 0.11 (0.05,0.26) | 0.11 (0.05,0.26) |
| | | NPIs & expanded vaccination | N/A | N/A | N/A | 0.02 (0.01,0.15) | 0.02 (0.01,0.14) | 0.02 (0.01,0.15) | 0.02 (0.01,0.15) |
| At-risk | Symptomatic (consult GP) | Expanded vaccination | 0.50 (0.47,0.57) | 0.58 (0.56,0.67) | 0.50 (0.47,0.57) | 0.50 (0.48,0.59) | 0.56 (0.54,0.64) | 0.57 (0.55,0.65) | 0.53 (0.50,0.60) |
| | | NPIs | 0.13 (0.06,0.34) | 0.29 (0.14,0.65) | 0.13 (0.06,0.34) | 0.11 (0.05,0.29) | 0.09 (0.04,0.24) | 0.09 (0.04,0.24) | 0.13 (0.06,0.33) |
| | | NPIs & expanded vaccination | 0.03 (0.01,0.16) | 0.07 (0.03,0.38) | 0.03 (0.01,0.17) | 0.02 (0.01,0.15) | 0.02 (0.01,0.14) | 0.02 (0.01,0.14) | 0.03 (0.01,0.18) |
| | Inpatient admissions | Expanded vaccination | 0.50 (0.47,0.57) | 0.59 (0.57,0.67) | 0.53 (0.50,0.59) | 0.52 (0.49,0.60) | 0.59 (0.56,0.66) | 0.60 (0.58,0.67) | 0.57 (0.54,0.64) |
| | | NPIs | 0.13 (0.06,0.34) | 0.29 (0.14,0.63) | 0.16 (0.07,0.38) | 0.13 (0.06,0.31) | 0.10 (0.05,0.26) | 0.11 (0.05,0.26) | 0.13 (0.06,0.32) |
| | | NPIs & expanded vaccination | 0.03 (0.01,0.16) | 0.06 (0.02,0.35) | 0.03 (0.01,0.18) | 0.02 (0.01,0.15) | 0.02 (0.01,0.15) | 0.02 (0.01,0.15) | 0.02 (0.01,0.17) |
| | In-hospital mortality | Expanded vaccination | 0.52 (0.50,0.58) | 0.60 (0.58,0.68) | 0.53 (0.50,0.59) | 0.52 (0.49,0.60) | 0.59 (0.56,0.66) | 0.60 (0.58,0.67) | 0.58 (0.56,0.65) |
| | | NPIs | 0.15 (0.07,0.36) | 0.30 (0.14,0.63) | 0.16 (0.07,0.39) | 0.13 (0.06,0.31) | 0.10 (0.05,0.26) | 0.11 (0.05,0.26) | 0.11 (0.05,0.28) |
| | | NPIs & expanded vaccination | 0.02 (0.01,0.17) | 0.06 (0.02,0.34) | 0.03 (0.01,0.19) | 0.02 (0.01,0.15) | 0.02 (0.01,0.15) | 0.02 (0.01,0.15) | 0.02 (0.01,0.15) |
| | Out of hospital mortality | Expanded vaccination | N/A | N/A | N/A | 0.52 (0.49,0.60) | 0.59 (0.57,0.66) | 0.60 (0.58,0.67) | 0.59 (0.57,0.66) |
| | | NPIs | N/A | N/A | N/A | 0.13 (0.06,0.31) | 0.10 (0.05,0.25) | 0.11 (0.05,0.26) | 0.11 (0.05,0.26) |
| | | NPIs & expanded vaccination | N/A | N/A | N/A | 0.02 (0.01,0.15) | 0.02 (0.01,0.14) | 0.02 (0.01,0.15) | 0.02 (0.01,0.15) |
| Low risk | Symptomatic (consult GP) | Expanded vaccination | 0.61 (0.58,0.69) | 0.63 (0.62,0.72) | 0.62 (0.59,0.70) | 0.40 (0.38,0.47) | 0.56 (0.54,0.64) | 0.57 (0.55,0.65) | 0.57 (0.54,0.65) |
| | | NPIs | 0.13 (0.06,0.34) | 0.28 (0.13,0.63) | 0.13 (0.06,0.35) | 0.11 (0.05,0.30) | 0.09 (0.04,0.24) | 0.09 (0.04,0.24) | 0.15 (0.07,0.40) |
| | | NPIs & expanded vaccination | 0.03 (0.01,0.20) | 0.08 (0.03,0.41) | 0.03 (0.01,0.21) | 0.02 (0.01,0.12) | 0.02 (0.01,0.14) | 0.02 (0.01,0.14) | 0.04 (0.01,0.23) |
| | Inpatient admissions | Expanded vaccination | 0.61 (0.58,0.69) | 0.63 (0.61,0.71) | 0.65 (0.62,0.72) | 0.42 (0.39,0.48) | 0.59 (0.56,0.66) | 0.60 (0.58,0.67) | 0.60 (0.57,0.67) |
| | | NPIs | 0.13 (0.06,0.34) | 0.29 (0.14,0.62) | 0.16 (0.07,0.37) | 0.14 (0.06,0.32) | 0.11 (0.05,0.26) | 0.11 (0.05,0.26) | 0.16 (0.07,0.39) |
| | | NPIs & expanded vaccination | 0.03 (0.01,0.20) | 0.07 (0.03,0.38) | 0.03 (0.01,0.22) | 0.02 (0.01,0.12) | 0.02 (0.01,0.15) | 0.02 (0.01,0.15) | 0.03 (0.01,0.22) |
| | In-hospital mortality | Expanded vaccination | 0.65 (0.61,0.72) | 0.71 (0.70,0.80) | 0.66 (0.62,0.72) | 0.41 (0.39,0.47) | 0.59 (0.57,0.66) | 0.60 (0.58,0.67) | 0.58 (0.55,0.65) |
| | | NPIs | 0.15 (0.07,0.37) | 0.40 (0.19,0.79) | 0.17 (0.08,0.39) | 0.13 (0.06,0.31) | 0.10 (0.05,0.25) | 0.11 (0.05,0.26) | 0.12 (0.06,0.30) |
| | | NPIs & expanded vaccination | 0.03 (0.01,0.22) | 0.09 (0.04,0.51) | 0.04 (0.01,0.24) | 0.02 (0.01,0.12) | 0.02 (0.01,0.14) | 0.02 (0.01,0.15) | 0.02 (0.01,0.17) |
| | Out of hospital mortality | Expanded vaccination | N/A | N/A | N/A | 0.41 (0.39,0.47) | 0.59 (0.57,0.66) | 0.60 (0.58,0.67) | 0.59 (0.56,0.65) |
| | | NPIs | N/A | N/A | N/A | 0.13 (0.06,0.31) | 0.10 (0.04,0.25) | 0.11 (0.05,0.26) | 0.11 (0.05,0.27) |
| | | NPIs & expanded vaccination | N/A | N/A | N/A | 0.02 (0.01,0.12) | 0.02 (0.01,0.14) | 0.02 (0.01,0.15) | 0.02 (0.01,0.14) |

**Table S4: For the pre-2021/2022 influenza season analysis, summary statistics for the age-stratified health episode occurrences as proportions relative to the non-COVID counterfactual scenario.** For the 100 simulations performed per scenario, we report the medians and in parentheses the 95% prediction intervals. We report estimates for each statistic to two decimal places. We assumed there were no out of hospital fatalities due to seasonal influenza for 0-49 year olds, with corresponding cells having value N/A.

| | | Age group (years) | | | | | | |
|---|---|---|---|---|---|---|---|---|
| **Risk status** | **Statistic** | **0-1** | **2-17** | **18-49** | **50-64** | **65-84** | **85+** | **All** |
| Overall population | Symptomatic (consult GP) | 1.17 (1.11,1.32) | 1.16 (1.12,1.27) | 1.17 (1.12,1.33) | 1.17 (1.12,1.34) | 1.17 (1.12,1.35) | 1.18 (1.12,1.35) | 1.17 (1.12,1.32) |
| | Inpatient admissions | 1.17 (1.11,1.32) | 1.28 (1.18,1.69) | 1.38 (1.22,2.17) | 1.38 (1.22,2.18) | 1.38 (1.21,2.18) | 1.38 (1.22,2.19) | 1.36 (1.21,2.03) |
| | In-hospital mortality | 1.37 (1.21,2.16) | 1.38 (1.22,2.12) | 1.37 (1.21,2.17) | 1.37 (1.21,2.18) | 1.38 (1.21,2.18) | 1.38 (1.22,2.19) | 1.38 (1.22,2.18) |
| | Out of hospital mortality | N/A | N/A | N/A | 1.37 (1.21,2.18) | 1.38 (1.22,2.18) | 1.38 (1.22,2.19) | 1.38 (1.22,2.18) |
| At-risk | Symptomatic (consult GP) | 1.17 (1.11,1.33) | 1.16 (1.11,1.27) | 1.17 (1.11,1.33) | 1.17 (1.12,1.33) | 1.17 (1.12,1.35) | 1.18 (1.12,1.35) | 1.17 (1.12,1.33) |
| | Inpatient admissions | 1.19 (1.13,1.40) | 1.28 (1.18,1.69) | 1.37 (1.21,2.17) | 1.37 (1.21,2.17) | 1.38 (1.21,2.18) | 1.38 (1.22,2.19) | 1.37 (1.21,2.12) |
| | In-hospital mortality | 1.37 (1.21,2.16) | 1.38 (1.22,2.13) | 1.37 (1.21,2.17) | 1.37 (1.21,2.17) | 1.38 (1.21,2.18) | 1.38 (1.22,2.19) | 1.38 (1.22,2.18) |
| | Out of hospital mortality | N/A | N/A | N/A | 1.37 (1.21,2.17) | 1.38 (1.22,2.18) | 1.38 (1.22,2.19) | 1.38 (1.22,2.18) |
| Low risk | Symptomatic (consult GP) | 1.17 (1.11,1.32) | 1.17 (1.12,1.27) | 1.18 (1.12,1.33) | 1.18 (1.12,1.34) | 1.17 (1.12,1.35) | 1.18 (1.12,1.35) | 1.17 (1.12,1.32) |
| | Inpatient admissions | 1.16 (1.11,1.31) | 1.28 (1.18,1.69) | 1.38 (1.22,2.18) | 1.38 (1.22,2.19) | 1.38 (1.22,2.18) | 1.38 (1.22,2.19) | 1.33 (1.20,1.86) |
| | In-hospital mortality | 1.37 (1.21,2.15) | 1.37 (1.22,2.08) | 1.38 (1.22,2.18) | 1.38 (1.22,2.19) | 1.38 (1.22,2.18) | 1.38 (1.22,2.19) | 1.38 (1.22,2.18) |
| | Out of hospital mortality | N/A | N/A | N/A | 1.38 (1.22,2.19) | 1.38 (1.22,2.18) | 1.38 (1.22,2.19) | 1.38 (1.22,2.19) |

### References

[1] Hill EM, Petrou S, Forster H, de Lusignan S, Yonova I, *et al.* Optimising age coverage of seasonal influenza vaccination in England: A mathematical and health economic evaluation. *PLOS Computational Biology* **16**(10):e1008278 (2020). doi:10.1371/journal.pcbi.1008278.

[2] Baguelin M, Flasche S, Camacho A, Demiris N, Miller E, *et al.* Assessing optimal target populations for influenza vaccination programmes: an evidence synthesis and modelling study. *PLoS medicine* **10**(10):e1001527 (2013).

[3] Hill EM, Petrou S, de Lusignan S, Yonova I, Keeling MJ. Seasonal influenza: Modelling approaches to capture immunity propagation. *PLOS Comput. Biol.* **15**(10):e1007096 (2019). doi:10.1371/journal.pcbi.1007096.

[4] Fumanelli L, Ajelli M, Manfredi P, Vespignani A, Merler S. Inferring the structure of social contacts from demographic data in the analysis of infectious diseases spread. *PLOS Computational Biology* **8**(9):e1002673 (2012). doi:10.1371/journal.pcbi.1002673.

[5] Office for National Statistics. Dataset: Population Estimates for UK, England and Wales, Scotland and Northern Ireland (2021). URL https://www.ons.gov.uk/peoplepopulationandcommunity/populationandmigration/populationestimates/datasets/populationestimatesforukenglandandwalesscotlandandnorthernireland. [Online] (Accessed: 13 October 2022).
